# IDENTIFYING PLAUSIBLE RANGES FOR DIFFERENTIAL VACCINE EFFICACY ACROSS HIGH- AND LOW-INCOME SETTINGS: A SYSTEMATIC REVIEW, DESCRIPTIVE META-ANALYSIS, AND ILLUSTRATIVE EVIDENCE ANALYSIS

**DOI:** 10.1101/2024.07.31.24310913

**Authors:** Esther Nyadzua Katama, Katherine E. Gallagher, Anoop Shah, D. James Nokes, David A McAllister

## Abstract

**Background:** Randomized clinical trials provide the highest standard of evidence about vaccine efficacy. Modelling exercises such as in evidence synthesis and health economic models where efficacy estimates are combined with other data to obtain effectiveness and cost-effectiveness estimates help inform policy decisions. The main challenge with such sensitivity analyses is in deciding on which assumptions to model.

**Purpose:** To identify plausible ranges for differential vaccine efficacy across high- and low-income settings.

**Data Sources and Study Selection:** MEDLINE, EMBASE, clinicaltrials.gov, and the World Health Organization International Clinical Trials Registry Platform (WHO-ICTRP) were searched for multi-site randomized clinical trials of bacterial and viral vaccines for the period of 01/01/1990 to 31/12/2020. Articles were restricted to those where at least one trial had included a low- or lower-middle-income setting, published in English, and conducted in humans.

**Methods:** A Bayesian random-effects meta-analysis was used to estimate the difference in vaccine efficacy in high-(high or upper middle) and low-(low or lower middle) income settings. A single hierarchical model that included all trials was used so that the degree to which estimates of vaccine efficacy against different diseases influenced one another was estimated from the observed data.

**Results:** Across 65 eligible trials (37 high-income, 21 low-income, and 7 both) covering 7 pathogens, only one trial reported efficacy estimates stratified by setting. Trials were similar in terms of design across settings. There was evidence of heterogeneity by vaccine target, typhoid vaccine demonstrated higher vaccine efficacy in low-income settings than in high-income settings but for all other vaccines, the point estimates indicated efficacy was lower in low-income settings; however, all credible intervals crossed the null.

**Conclusions:** The percentage of trials in low-income settings poorly reflects the burden of disease experienced in low-income settings. While there is evidence of lower vaccine efficacy in low-income settings relative to high-income settings, the credible intervals were very wide. Vaccine efficacy trials should report treatment effects stratified by settings.

## Introduction

Vaccination is important for reducing the burden of life-threatening infectious diseases, preventing between 2 and 3 million deaths every year^1,2^. Therefore, policy decisions concerning the choice of vaccines and whom to vaccinate are crucial for public health. For guiding such decisions, randomised clinical trials (hereafter trials) provide the highest standard of evidence about vaccine efficacy.

Efficacy estimates (usually reported on the relative scale, e.g., hazard ratios, relative risk reductions) alone, however, are insufficient to guide policy. Policymakers also need to decide whether vaccines are likely to be effective (usually reported as an absolute effect such as a risk difference) and cost-effective (e.g., the cost per quality-adjusted life year) in real-world settings. Such decision-making can be supported by modelling exercises such as in evidence synthesis and health economic models where efficacy estimates are combined with other data (such as rates of infection in low-income settings) to obtain effectiveness and cost-effectiveness estimates respectively.

Such modelling requires explicit assumptions about the applicability of relative effects (derived from trials) to real-world settings. However, when considering low-income settings this assumption is more difficult because, despite low-income settings having a higher burden of communicable diseases^3,4^, most vaccine trials are conducted in high-income settings. Additionally, there are biologically plausible factors/reasons (such as genetics, sex, and comorbidities) why efficacy might differ by income setting.^5^

Nonetheless, even where low-income trials are lacking, it is still possible to model effectiveness and cost-effectiveness. Modellers can conduct “sensitivity analyses” under different assumptions about relative efficacy; they can obtain estimates under different assumptions as to the difference in efficacy between income settings. Depending on the circumstances, such sensitivity analyses might provide reassurance; alternatively, they can identify where effectiveness and cost-effectiveness is most uncertain, helping prioritise where additional evidence from low-income settings would be most beneficial.

The main challenge with such sensitivity analyses is in deciding on which assumptions to model due to expert elicitation. These decisions should be informed by the deliberations of scientists with appropriate expertise.^5,6,7^ Their deliberations, however, can benefit from relevant empirical evidence.^8^ Empirical estimates of difference in efficacy across settings from other vaccine trials provide a plausible range to help inform their advice. For example, if large differences in efficacy between low- and high-income settings are common across vaccine trials, this would lead to different sensitivity analyses than if such difference were rare.

Therefore, to inform such models, we conducted a systematic review and meta-analysis for different vaccines, comparing efficacy across different income settings.

## Methods

### Systematic search strategy and criteria

We searched MEDLINE/PubMed, EMBASE, clinicaltrials.gov (https://clinicaltrials.gov/) and the World Health Organization International Clinical Trials Registry Platform (WHO-ICTRP - http://apps.who.int/trialsearch/) for randomized clinical trials conducted from January 1990 to December 2020. Publications were identified using pathogen specific, pathogen vaccine specific search terms and Boolean operators. The following search terms were used: “anthrax”, “brucellosis”, “cholera”, “diphtheria”, “haemophilus influenzae b”, “meningococcal”, “pertussis”, “plague”, “ pneumococcal”, “tetanus”, “tuberculosis”, “typhoid”, “typhus”, “exanthematicus”, “Encephalitis”, “Influenza”, “Hepatitis”, “Measles”, “Mumps”, “Poliomyelitis”, “Rabies”, “Rotavirus diarrhea”, “Rubella”, “Varicella zoster”, “Yellow fever”, “Papillomavirus”, “vaccin*”, “immuni*”, “human” and “inoculat*”. Our search was restricted to randomized controls trial articles published in English published between 1990 and 2020 and to studies conducted in humans. A PRISMA flow diagram outlined the study selection process (figure 1)^9^. Eligibility of abstracts and full texts was assessed by three reviewers based on the inclusion criteria. Any discrepancies or disagreements were resolved by consensus. We contacted the study authors of trials conducted in multiple countries that did not report vaccine efficacy data stratified by country.

**Figure 1.**
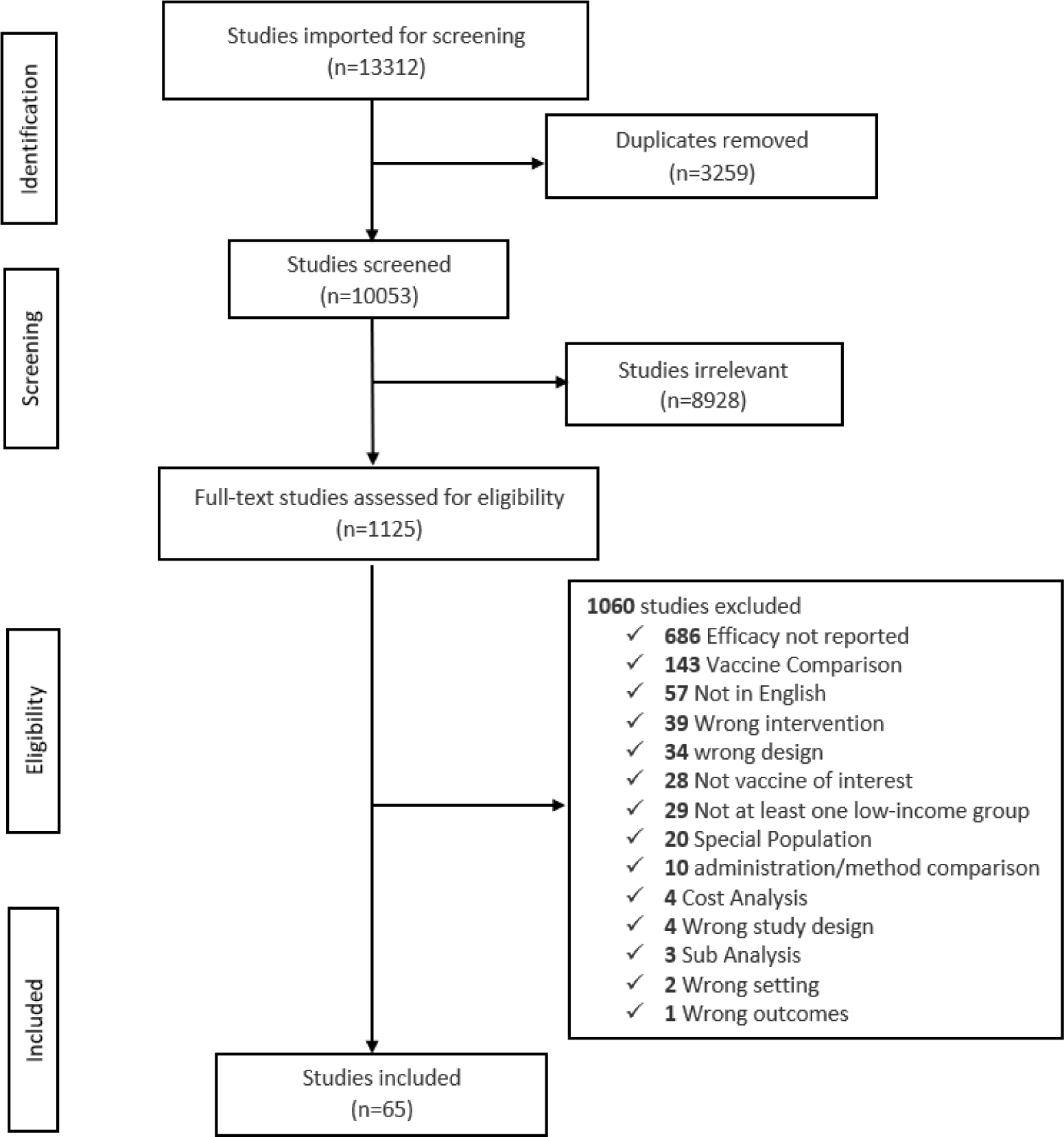
PRISMA flowchart outlining the study selection process.

### Inclusion criteria

We aimed to capture articles including a “vaccine of interest” for all ages (Bacterial: anthrax vaccines, brucellosis vaccines, cholera vaccines, diphtheria vaccines, *Haemophilus influenzae* type b vaccines, meningococcal vaccines, pertussis vaccines, plague vaccines, pneumococcal vaccines, tetanus vaccines, tuberculosis vaccines, typhoid vaccines, typhus (exanthematicus) vaccines; Viral: encephalitis vaccines, influenza vaccines, hepatitis vaccines, measles vaccines, mumps vaccines, poliomyelitis vaccines, rabies vaccines, rotavirus vaccines, rubella vaccines, Varicella-zoster vaccines, yellow fever vaccines, human papillomavirus vaccines) where randomization was performed at the level of individual patient. Trials were limited to phase two, three and four.

### Exclusion criteria

We excluded any trial without a report in English and trials that did not have at least one site in a low- or lower-middle income country. We also excluded trials in special populations including people with HIV, chronic illness, and premature babies. We excluded abstracts where the full text was not available if the authors did not respond to requests for results. After obtaining summary data for each trial, we also excluded high-income trials for older age groups (1 trial from pneumococcal trials, 7 influenza trials and 1 hepatitis trial) because there were no comparable trials from low-income settings.

### Outcome of Interest

The primary outcome was vaccine efficacy against disease and incident infection (all trials required symptomatic infection, most also required a positive test for the relevant pathogen). Where multiple effect estimates were presented for incident infection (e.g., type-specific and non-type specific), the primary outcome was selected.

### Exposure

Each trial site was assigned an economic status using the World Bank classification^10,11^. Each trial was classified according to the site of the trial into low-, lower middle-, upper middle- or high-income. Multisite trials across different income levels were classified as “both”. These income or exposure definitions were finalised prior to plotting trial effect estimates or fitting regression models.

### Covariates

We defined the following covariates based on trial reports – phase, blinding, type of control arm, study year, mode of administration, type of analysis, and type of vaccine.

### Data Extraction

Covidence software (https://www.covidence.org/) was used for article screening and full text review. Effect estimates and trial-level characteristics were extracted for each trial. Data on study design, sample size, country, vaccine type, age groups, duration of study, method of administration, reported vaccine efficacy and 95% confidence intervals (CIs) were also collected. Extracted data were entered into a data collection form created in Microsoft Excel.

### Assessing representativeness of trial populations

Generally, low-income countries have a higher burden of disease than countries in higher income categories. These countries are often under-represented^3^ in research which might bias the overall (average) efficacy. To understand this bias, we compared the geographic representation of participants in clinical trials compared to the geographic variation in the burden of disease ^12–24^. The percentage of trial participants from each setting was compared to the prevalence of disease in each setting (figure 2).

**Figure 2.**
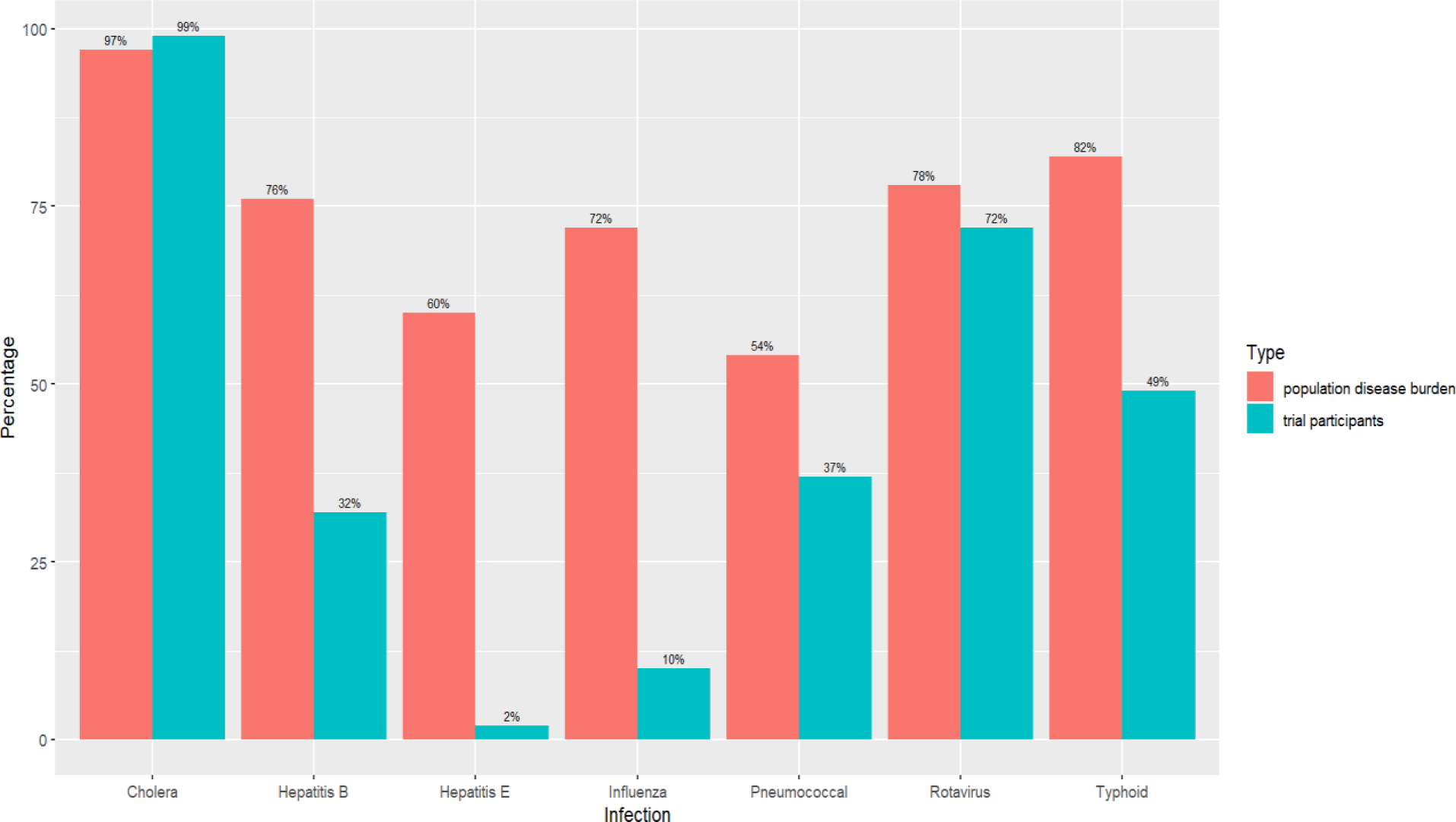
Comparison of the geographic representation of participants in clinical trials compared to the geographic variation in the burden of disease in Low-Income Countries. The percentage of trial participants from each setting was compared to the prevalence of disease in each setting. The blue bars indicate the total trial participants in LICs. Red bars indicate the total population in LICs.^1^

### Data management and statistical Analysis

Trial characteristics were cross tabulated by trial income setting. Trial relative effect estimates (usually percentage reduction) were transformed into log-ratios (log-odds ratio, log-hazard ratios or log rate ratios depending on the measure reported in the trial reports) hereafter termed log-RRs. We log-transformed the point estimates and upper and lower limits of the 95% confidence intervals. The standard errors were then estimated from the transformed 95% confidence interval limits (standard error = (upper limit – lower limit) / (2*1.96)). For each infection, we plotted the effect estimates and 95% confidence intervals for low-income, high-income and both.

Trials from both high- and low-income settings were excluded from further analyses. We then fitted Bayesian hierarchical models to estimate the difference in vaccine efficacy in high- and low-income settings. We used Bayesian models as these allowed partial pooling between trials for different pathogens. This meant that separate estimates for difference in efficacy by income setting could be obtained for each infection, while allowing estimates for different pathogens to influence each other. We used hierarchical models so that the degree to which estimates from different pathogens influenced each other was estimated from the observed data. Rather than separately analysing trials for each pathogen, we included all trials in a single hierarchical model for two reasons. First because this approach shrinks any extreme values towards the overall average and secondly because it improves precision due to greater sample size.

The models included a normal likelihood (y ∼ N (μ, s)) where y was the log-RR and s was the SE_log-RR_ from the trial reports. The linear predictor for μ included group terms for both trial and infection. Nested within each group there was an intercept term (for the efficacy in high income trials) and a term for the low-income versus high income comparison (coded 0 for high income and 1 for low income). Thus, after model fitting, the efficacy in low-income settings could be obtained by summing samples from the intercept and comparison. All models also included age of trial participants. To account for the distributions of ages within each trial, we sampled from the mean age and standard deviation within the model. For all three quantities of interest (high income, high income versus low income and low income) estimates of the central tendency were obtained from the mean of the samples from the model and upper and lower credible intervals were obtained as the 2.5^th^ and 97.5^th^ percentiles. These were then exponentiated to the ratio scale (RRs). Finally, in the text we converted the RRs into relative risk reductions (RRR = 100 x (1-RR)) as this is the most conventional estimate reported in the vaccine literature We retained (log) RRs in the figures as the interactions.

In additional exploratory models, we included terms for phase, year of trial, blinding, type of control arm, mode of administration, type of analysis, age, and type of vaccine to examine variation in efficacy by income setting after adjusting for any other known differences between trials. Due to the relatively small amount of data, we did not attempt to simultaneously model all these covariates, instead including each in turn.

We also performed an illustrative evidence synthesis showing how the interaction estimates (i.e., the difference in efficacy from low and high-income settings) could be used to predict efficacy in low-income settings. We did so for a range of scenarios where a vaccine has strong, moderate, or weak efficacy in high income settings (RR 0.2, 0.5 and 0.8 respectively) and where these efficacies had been estimated with low, intermediate, and high precision (corresponding to standard errors of the log-RR of 0.50, 0.25 and 0.10 respectively). We produced uncertainties estimates for the predictions as follows. First, we assumed that the log-RRs for the high-income scenarios were normally distributed and for each scenario obtained i) 6,000 samples from normal distribution (e.g., high efficacy low precision mean = log (0.2), SD = 0.50). For the interaction efficacy estimates we obtained ii) 6,000 samples directly from the output of the Bayesian models (3 chains of 2,000 samples each). For this interaction estimate we obtained predictions for some notional unobserved vaccine (i.e., one not included in our meta-analysis) by sampling (with equal probability) from the posteriors for each of the seven observed vaccines. For each of the 6,000 samples, a predicted low-income setting estimate was then obtained by summing i) and ii). The distributions were then presented as density plots. The probability that the low-income efficacy was better than 1.0, 0.8, 0.7 and 0.5 were also calculated by calculating the proportion of samples below each level (e.g., for RR 0.7, number of samples where log (RR) < log (0.7)/total number of samples). Full descriptions of the model, the extracted data, and model code are provided in the supplementary appendix. All analyses were performed in R. Bayesian models were fitted using the BRMS package^25^, which fits Bayesian models using Hamiltonian Markov Chain Monte Carlo (MCMC).

## Results

13312 publications met our search criteria with 65 trials^26–118^ meeting our inclusion criteria (figure 1). All the 65 studies that met our inclusion criteria reported vaccine efficacy of one of the vaccines of interest.

### Trial characteristics

Most of the trials were high-income (57%). The majority were placebo-controlled, double blind and Phase III (table 1). 32% of the eligible trials were from low-income countries, in Phase III and double-blind. There were not any single-blind trials conducted in low-income countries. Trials were similar in terms of design across settings (e.g., phase 3 86% and 76%, double blind: 73% and 71%, and vaccine 35% and 33% for high versus low respectively). For pneumococcal vaccines, trials in low-income countries were generally of lower valency and trials in high income countries tested higher valency vaccines. Trials in high-income countries (HICs) were pneumococcal 7-valent conjugate vaccine (PCV7), pneumococcal conjugate vaccine (PCV13), 14-valent pneumococcal conjugate vaccine (PCV14) and the trials in low-income countries (LICs) were 9-valent pneumococcal conjugate vaccine (PCV9), 11-valent pneumococcal conjugate vaccine (PCV11) (neither of which were licenced in the end) and 10-valent pneumococcal conjugate vaccine (PCV10). For rotavirus vaccines, the type of coverage was similar for trials in both high-income countries and low-income countries (Supplementary, table 1).

**Table 1.** Trial-level characteristics of eligible randomized control trials such as study design, sample size, country, vaccine type, and method of administration. Countries are stratified as high and low income or both.

|  |  | Income n(%) |  |  | Overall |
| --- | --- | --- | --- | --- | --- |
|  |  | High | Both high & low | Low |  |
| Participants | N = 891959 | 471257 (53%) | 54906 (6%) | 365795 (41%) | 891959 |
| Trials | N = 65 | 37 (57%) | 7 (11%) | 21 (32%) | 65 |
| Pathogens | Hepatitis E | 1 (50%) | 0 (0%) | 1 (50%) | 2 |
|  | Cholera | 1 (50%) | 0 (0%) | 1 (50%) | 2 |
|  | Pneumococcal | 7 (70%) | 0 (0%) | 3 (30%) | 10 |
|  | Typhoid | 2 (29%) | 0 (0%) | 5 (71%) | 7 |
|  | Influenza | 17 (61%) | 6 (21%) | 5 (18%) | 28 |
|  | Hepatitis B | 2 (67%) | 0 (0%) | 1 (33%) | 3 |
|  | Rotavirus | 7 (54%) | 1 (8%) | 5 (38%) | 13 |
| Phase | Phase II | 2 (5%) | 0 (0%) | 2 (10%) | 4 |
|  | Phase III | 32 (86%) | 7 (100%) | 16 (76%) | 55 |
|  | Phase IV | 3 (8%) | 0 (0%) | 3 (14%) | 6 |
| Blinding | double-blind | 27 (73%) | 7 (100%) | 15 (71%) | 49 |
|  | open label | 6 (16%) | 0 (0%) | 6 (29%) | 12 |
|  | single-blind | 4 (11%) | 0 (0%) | 0 (0%) | 4 |
| Control | active-controlled trial | 15 (41%) | 4 (57%) | 8 (38%) | 27 |
|  | placebo-controlled | 19 (51%) | 3 (43%) | 11 (52%) | 33 |
|  | usual care | 3 (8%) | 0 (0%) | 2 (10%) | 5 |
| Type of vaccine | Inactivated Vaccines | 9 (24%) | 5 (71%) | 4 (19%) | 18 |
|  | Live-attenuated | 13 (35%) | 2 (29%) | 7 (33%) | 22 |
|  | not specified | 3 (8%) | 0 (0%) | 1 (5%) | 4 |
|  | recombinant \| polysaccharide \| conjugate | 12 (32%) | 0 (0%) | 9 (43%) | 21 |
| Mode of administration | Intramuscular | 17 (46%) | 1 (14%) | 11 (52%) | 29 |
|  | Mucosal | 12 (32%) | 1 (14%) | 6 (29%) | 19 |
|  | not specified | 8 (22%) | 5 (71%) | 4 (19%) | 17 |
| Type of analysis | intention-to-treat (ITT) | 3 (8%) | 0 (0%) | 4 (19%) | 7 |
|  | not specified | 16 (43%) | 0 (0%) | 7 (33%) | 23 |
|  | Per-protocol | 18 (49%) | 7 (100%) | 10 (48%) | 35 |

### Trial participants compared to real-world patients

The estimated population in low-income and lower-middle-income countries was 4.4 billion and in upper-middle income and high-income countries as 3.7 billion^12^. Cholera was estimated as 2.8 million cases in low-income setting^13^, pneumococcal as 2477 million cases^14,15^, typhoid as 17.8 million^16,17^, hepatitis B as 572 million cases^18^, rotavirus as 200 million episodes of diarrhea^19^, 65 million cases of influenza^20–23^ and hepatitis E as 797,764 cases in low-income setting^24^.

The percentage of trial participants living in low-income settings was in some instances considerably lower than the percentage of patients with disease living in low-income settings. For example, even though approximately 72% of cases of influenza occur in low-income countries^22,23^, only 10% of trial participants were in low-income countries. Even though approximately 60% of cases of Hepatitis E estimates occur in low-income countries^24^, only 2% of trial participants were in low-income countries (Figure 2).

### Efficacy in high- and low-income settings

On comparing the reported (i.e., not modelled) efficacy estimates across low income, high income and mixed income trials, the efficacy appeared to be greater in high income settings, although the 95% credible intervals overlapped (Supplementary, figure 1).

Across all pathogens, the point estimate for the vaccine efficacy was higher in high-income countries than low-income countries (i.e., pooled vaccine efficacy (VE) in HICs was 77% (95% CI 39% to 92%) compared to 65% (95% CI −9% to 89%) in LICs) (Figure 3). The RR for the interaction was 1.5 (95% CI 0.65 to 3.68) indicating that the most likely difference in efficacy is a 33% lower efficacy in low-income settings compared to high income settings. There was evidence of heterogeneity by vaccine target, typhoid vaccine demonstrated higher vaccine efficacy in low-income settings than in high-income settings but for all other vaccines, the point estimates indicated efficacy was lower in low-income settings; however, all credible intervals crossed the null. The difference varied from VE of 79% (95%CI) for influenza to VE of 30% for cholera vaccines. For all the comparisons, both for each infection and overall, the 95% credible intervals included the null, compatible with no difference in efficacy between low-income settings and high-income settings.

**Figure 3.**
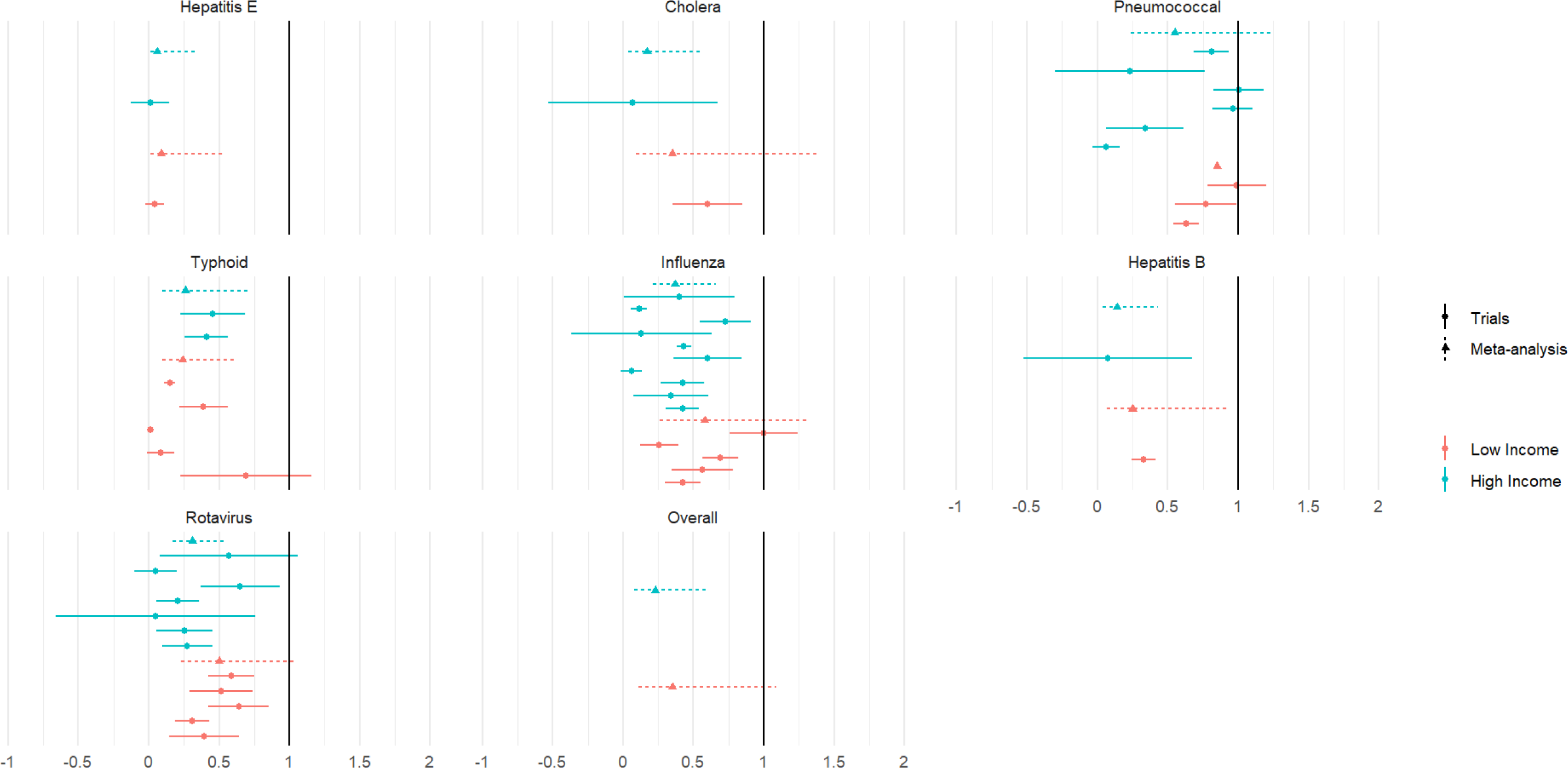
Comparison of trial effect estimates and the meta-analysis estimates in high- and low-income settings by infection and overall. The blue lines indicate the trials in HICs. Red lines indicate the trials in LICs. Dotted lines represent the meta-analysis estimates while the continuous line show the reported trial effect estimates.

In the models adjusting for the type of control arm, study year, phase, mode of administration, analysis (ITT versus unspecified), type of vaccine (Inactivated, Live attenuated, Recombinant) and blinding, the difference in efficacy estimates were similar across all models (Supplementary, table 3). Incidence outcomes were similar for low- and high-income trials (Supplementary, table 4). In a sensitivity analysis where we excluded 7 trials where the outcomes appeared to differ between high-income and low-income settings results were also similar (Supplementary Table 5) analysis.

### Illustrative evidence synthesis

Figure 4 illustrates the effect of combining i) the interaction estimates from our models comparing vaccine efficacy in low-income versus high-income settings (green density plots) with ii) notional efficacy estimates obtained from high income settings (pink density plots). Combining i) and ii) leads to iii) a predicted efficacy in low-income settings (blue density plots). In a formal evidence synthesis or cost-effectiveness analysis this would then be combined with other data (e.g., baseline rates of infection), but for simplicity these are not included in this illustration. The panels show these predictions for a range of different high-income scenarios, from strong to weak efficacy and from low to high precision. Since the low-income efficacy predictions incorporate the uncertainty from both i) and ii) they can be used to obtain estimates of probabilities for any low-income setting efficacy. As an example, taking the moderate efficacy/intermediate precision scenario, the probability of a low-income setting efficacy of better than no efficacy is 68%. The probabilities that the efficacy is better than20%, 30% and 50% is 53%, 44% and 23% respectively.

**Figure 4.**
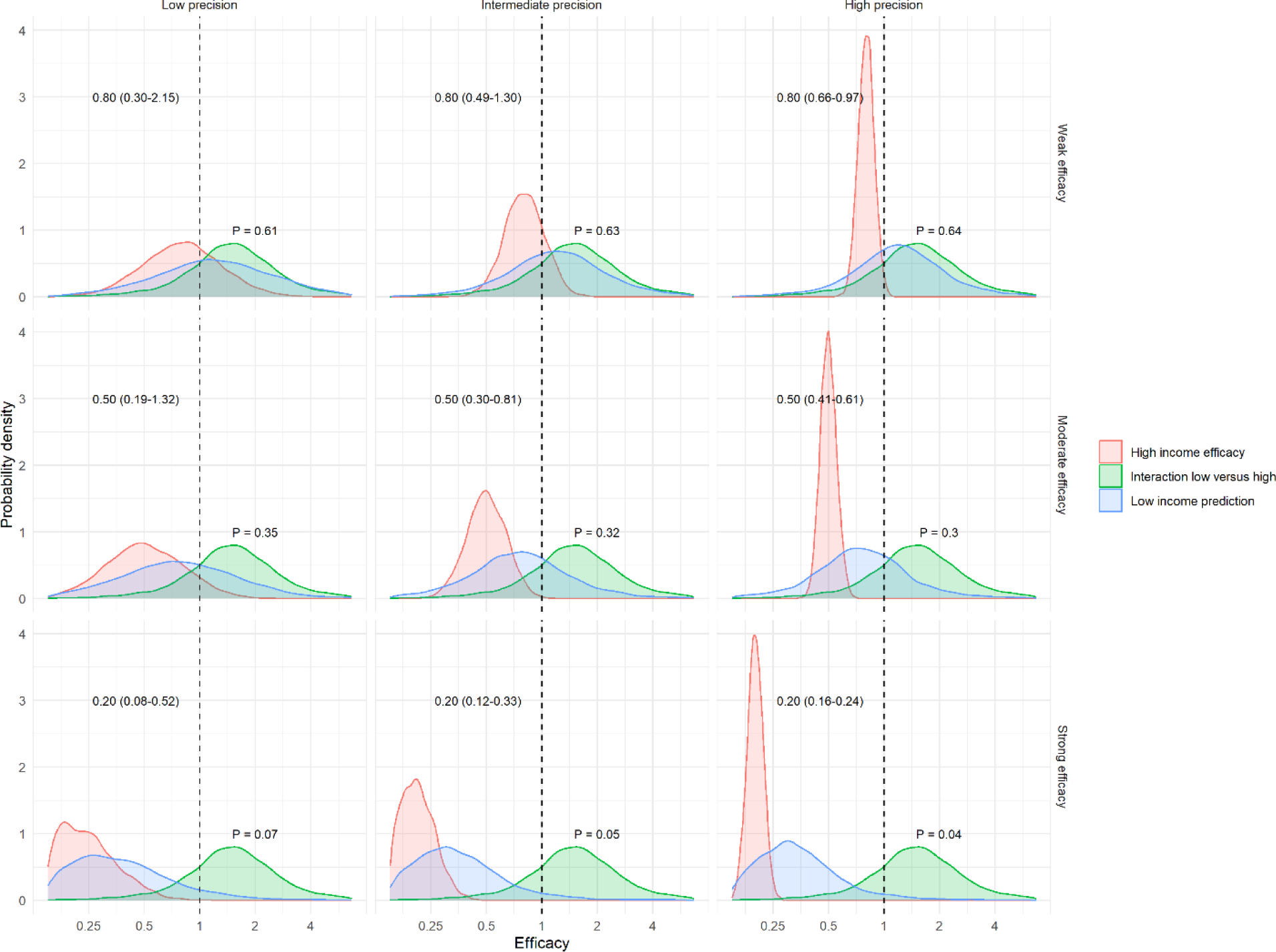
Illustration of vaccine efficacy in low-income settings under scenarios where the vaccine efficacy in high-income countries differ. The x-axis is modelled efficacy plotted on (log) rate ratio scale with labels on rate ratio scale. Red represents the high-income efficacy, green indicates the interaction between LICs and HICs while blue represents the low-income setting prediction.

Figure 5 shows a similar evidence synthesis, here the empirically derived interaction estimates have been modified, as if also informed by relevant scientific understanding. Again, taking the moderate efficacy/intermediate precision high-income scenario example, if the attenuated efficacy is believed to be more likely, the probability that the low-income efficacy is 30% or better is 12%. In contrast, where attenuated efficacy is believed less likely, the probability that the low-income efficacy is 30% or better is 77%.

**Figure 5.**
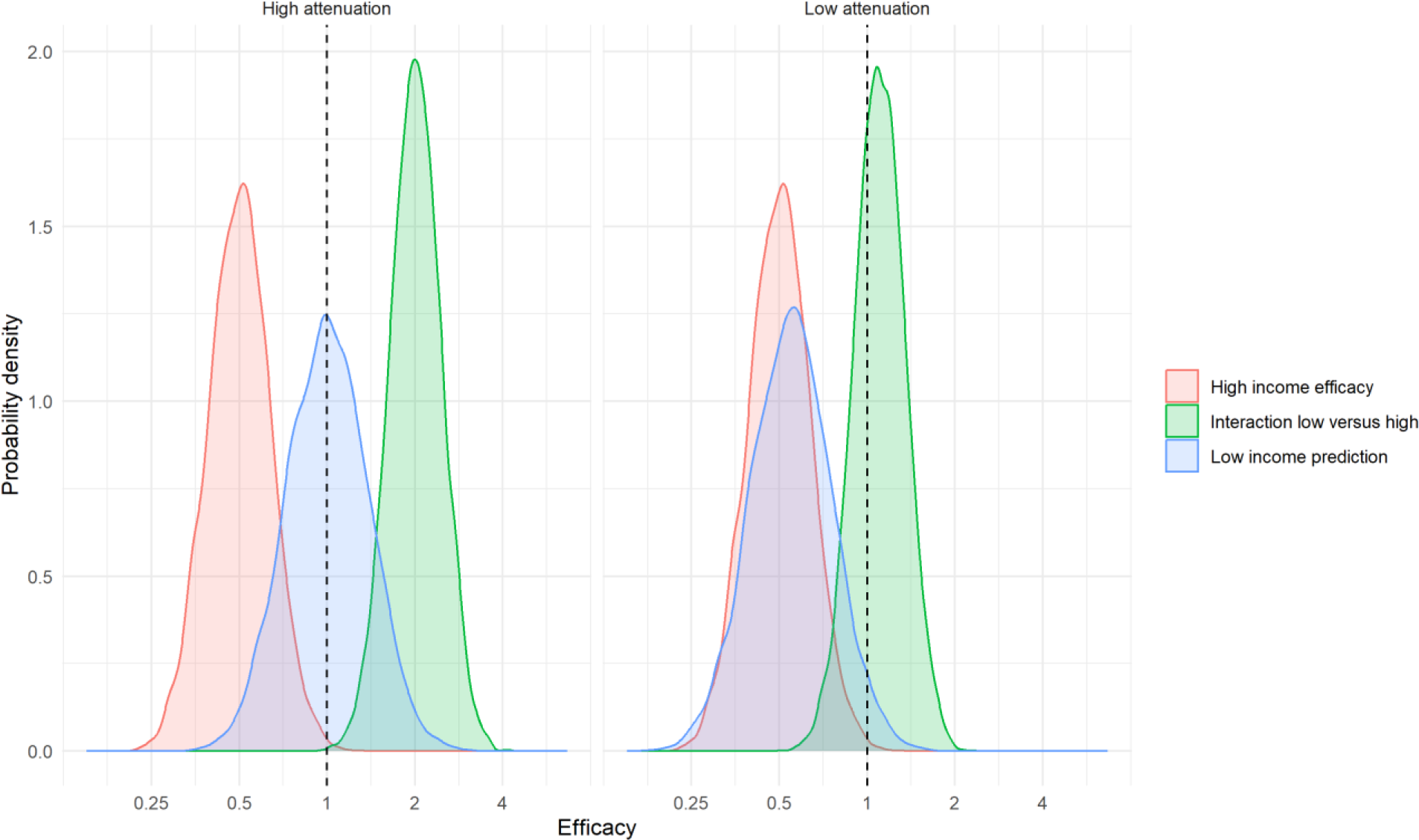
Illustration of impact of differential efficacy where interaction estimates are modified based on relevant expertise, e.g., mechanistic knowledge of likely differences in efficacy. The x-axis is modelled effect efficacy plotted on (log) rate ratio scale with labels on rate ratio scale. Red represents the high-income efficacy, green indicates the interaction between LICs and HICs while blue represents the low-income setting prediction.

## Discussion

### Principal findings

In a systematic review and meta-analysis of 65 trials across 7 pathogens we found that vaccine efficacy was on average around 33% lower in low income compared to high income settings with a high level of uncertainty ranging from higher to lower efficacy in low-income settings. In illustrative evidence synthesis we show how this finding can be combined with high-income efficacy estimates to predict vaccine efficacy in low-income settings, producing probability estimates for different levels of efficacy in in low-income settings.

Where evidence for low-income settings is absent for a specific vaccine, our findings, from across seven pathogens, can be used to inform expert deliberations as to the likely attenuation in efficacy; the results of such deliberation can then be used to inform evidence synthesis and cost-effectiveness models. Depending on the findings, such modelling may provide reassurance; alternatively, they may identify where effectiveness and cost-effectiveness is most uncertain, helping prioritise where additional evidence from low-income settings would be most beneficial.

While our comparisons were essentially descriptive and agnostic to the reasons for differences in efficacy between high- and low-income settings, it is useful to consider why we observed difference in efficacy of the magnitude seen in this meta-analysis. First, intrinsic host factors such as age at time of vaccination, sex and comorbidities, strongly influence vaccine responses to individuals hence difference in vaccine efficacy^5^. Secondly, there may be differences in environmental factors across regions where the vaccine is targeted for use in addition to disease prevalence and burden of disease^7^. Factors influencing the force of infection such as geographical location, rural versus urban environment, family size and season^5^, are some of the potential mechanisms for attenuation of efficacy. We observed a slight attenuation of efficacy in low versus high-income settings.

### Strengths and limitations of the study

Our review has compared efficacy in high versus low-income settings across multiple vaccines. We are not aware of other reviews of this nature. Our study does have several limitations. Trials were limited to English language which might have resulted in the exclusion of eligible studies. Also, we had a limited number of trials included in our analysis due to the few trials available from low-income countries compared to high-income countries.

The trials themselves were mostly exclusively in either high- or low-income settings, and for the few that were conducted in both settings we were unable to obtain results stratified by setting. Thus, setting comparisons were also between-trial comparisons. Consequently, apparent differences between settings could be confounded by differences between trials. Finally, while we compared a range of trial characteristics across low- and high-income settings, finding them largely similar, and adjusted for each of a number of characteristics in single-covariate models (e.g., phase, year of study), it remains possible that differences in trial design, completeness of outcome ascertainment etc, may cause some *apparent* attenuation in efficacy in low-income settings compared to high-income.

### Comparison with other studies

An observer-blind trial that reported on efficacy of influenza vaccine across diverse geographic regions reported that influenza vaccine efficacy was higher in high-income countries than in low-income countries. This finding is supported by a study review from Lesham et al, which indicated high efficacy for rotavirus strain-specific in high-income countries^119^. A review on performance of rotavirus vaccines in developed and developing countries reported a high efficacy in industrialized nations compared to poor settings despite the greatest rotavirus disease burden which is consistent with our findings^120,121^. Our own findings are generally consistent with these observations, adding empirical estimates from across a range of vaccines.

### Implications for practice and policy

We and others have found that people from low-income countries were under-represented in vaccine trials. This is true despite low-income countries bearing the greatest burden of disease. Reasons for this under-representation may include lack of research capacity due to lack of access to funding, ethical and regulatory system obstacles, limited financial and human capacity and operational barriers^3,122^ These factors lead to under-representation despite the disproportionate burden borne by the low-income countries. Pharmaceutical companies and manufacturers may be less likely to conduct trials in low-income settings due to a perceived risk of higher adverse event rates alongside the potential for major reductions in lower financial returns^122^. Consequently, policymakers in low-income countries are forced to make inferences based on vaccine efficacy data from higher income countries when determining vaccine policy.

While additional trials in low-income settings would be the best approach for guiding vaccine policymaking, in the absence of such trials, we have produced average estimates of the extent to which vaccines are, in general, likely to be less efficacious in low-income settings. In our illustrative evidence synthesis, we show how empirical evidence as to differential efficacy can be combined with estimates of efficacy from high-income settings to predict efficacy in low-income settings. We also show how our empirical interaction estimates could be treated as a starting point, to be updated based on scientific knowledge about the likelihood of differences in efficacy given the biology of specific vaccines.

Finally, we identified an issue with trial reporting. Even where trials included low-income settings, the disaggregated effect estimates were rarely reported for these settings. Had this been available we would have been able to produce more precise and robust estimates of heterogeneity in treatment effects across settings. We suggest that clinical trials of vaccines should routinely report with treatment effects stratified by income settings, as such estimates can be used to inform meta-analyses, even where there is insufficient data to draw conclusions from single trials. Such information would lead to improved interaction estimates for the difference in efficacy from across settings. This would allow evidence synthesis to be conducted with fewer assumptions, with greater confidence in supporting decision-making.

## Conclusion

We found that the percentage of trial participants in low-income settings was lower than the proportion of individuals with the relevant disease burden in low-income settings. We found that there was some evidence (with a wide uncertainty range including the null) for a modest attenuation of efficacy in low-versus high-income settings and show how the uncertainty around this estimate can be combined with efficacy results from high-income settings to predict efficacy in low-income settings.^123^ While it is frequently claimed that heterogeneity in treatment effects (HTE) is uncommon in clinical trials, evidence for this for vaccines across settings has not previously been presented. We provide support for this general statement. Therefore, although there may be theoretical reasons why this is not true for specific vaccines, our observations provide a good starting point for discussing the potential of HTE by region.

## Data Availability

All data produced are available online at
https://zenodo.org/record/7956407

## Acknowledgements

The authors thank Mr Alex Maina for his advice and guidance concerning the search of the trials reported within this article.

## Footnotes

1 The blue bars indicate the total trial participants in LICs. Red bars indicate the total population in LICs.

