## Supplementary File for "IDENTIFYING PLAUSIBLE RANGES FOR DIFFERENTIAL VACCINE EFFICACY ACROSS HIGH- AND LOW-INCOME SETTINGS: A SYSTEMATIC REVIEW, DESCRIPTIVE META-ANALYSIS, AND ILLUSTRATIVE EVIDENCE ANALYSIS"

### Supplementary Figure 1: Forest plot of reported trial effect estimates by income category for each infection

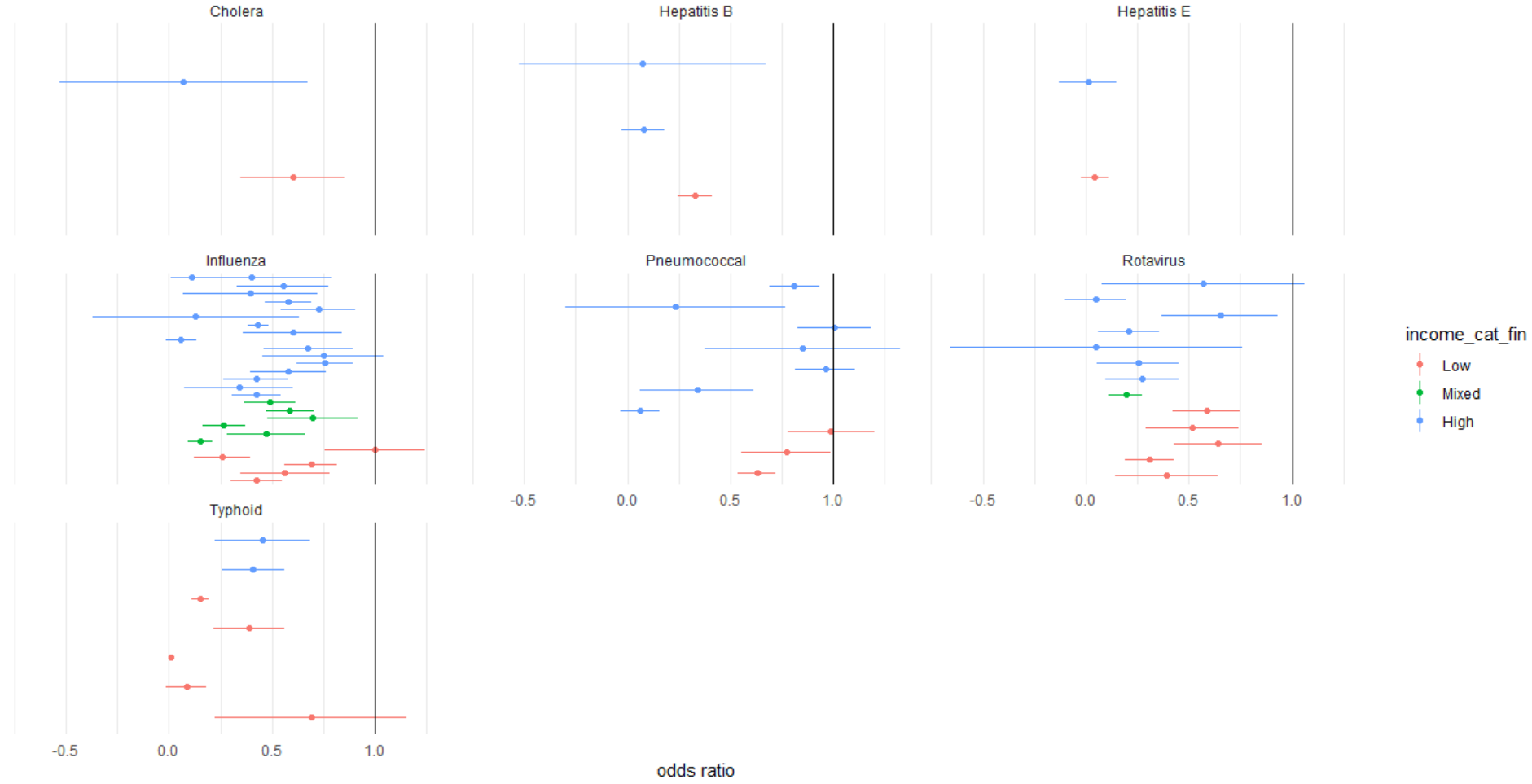

### Supplementary Table 1: Rotavirus and Pneumococcal serotypes

| **Rotavirus** | | |
| --- | --- | --- |
| **Vaccine serotype** | **High-Income trials (n)** | **Low-Income trials (n)** |
| RotaTeq (Merck) | 1 | 1 |
| RotaSIIL (BRV-PV, Serum Institute of India) | 2 | 2 |
| Rotarix (GlaxoSmithKline Biologicals) | 2 | 1 |
| RotaShield (Wyeth; IDT Biologika GmbH) | 1 | 1 |
| RIT 4237 | 1 | 0 |
| **Pneumococcal** | | |
| **Vaccine serotype** | **High-Income trials (n)** | **Low-Income trials (n)** |
| PCV7 (Wyeth) | 4 | 0 |
| PCV9 (Wyeth) | 0 | 1 |
| PCV10 (Synflorix; GSK) | 0 | 1 |
| PCV11 (sanofi pasteur) | 0 | 1 |
| PCV13 (Pfizer) | 2 | 0 |
| PCV14 (Merck) | 1 | 0 |

### Supplementary Table 2: Efficacy in high- and low-income settings and the difference between each, by infection and overall

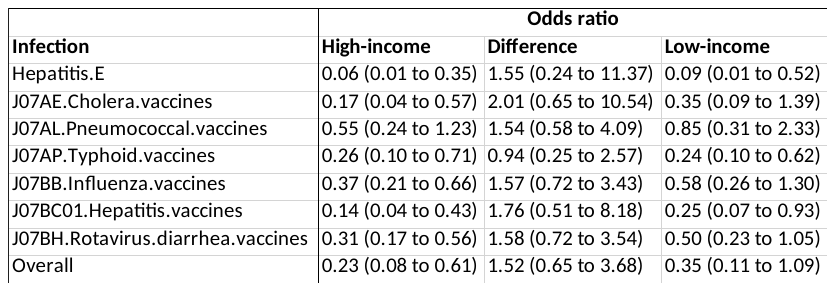

### Supplementary Table 3: Model Effect Estimates

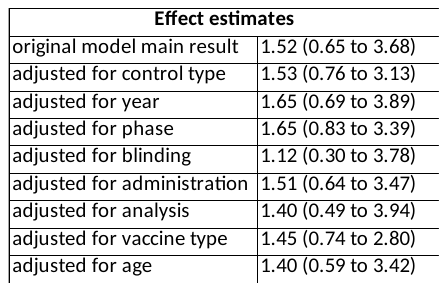

### Supplementary Table 4: Outcomes^[1]^

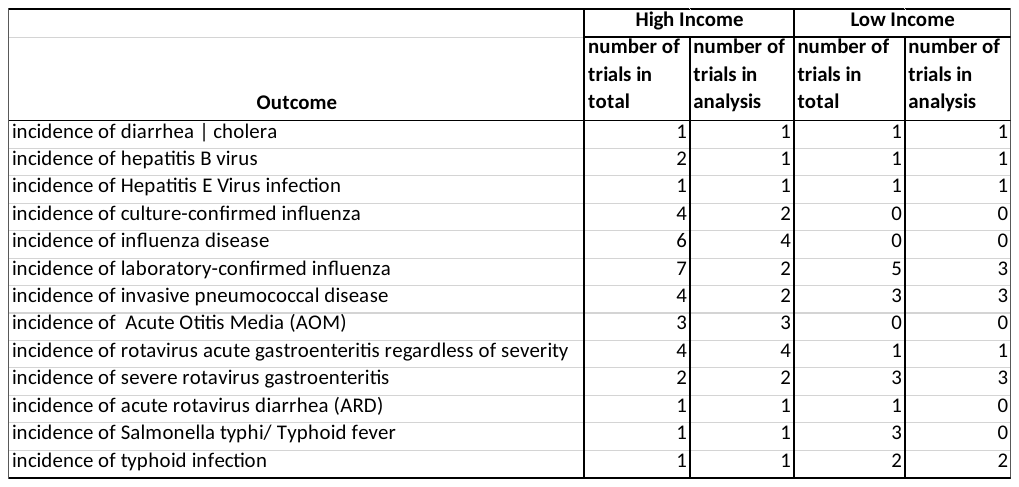

### Supplementary Table 5: Sensitivity Analysis

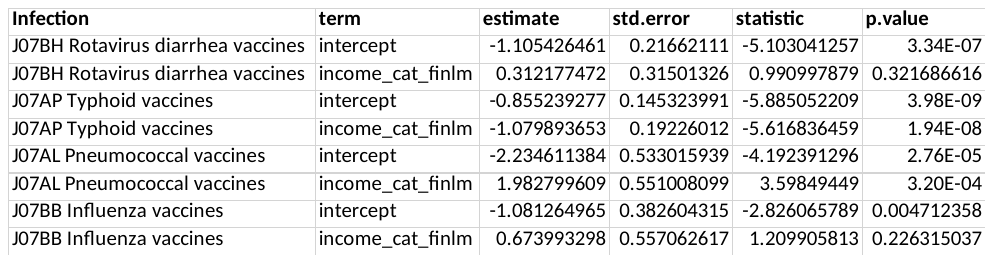

1. Incidence similar for trials
